# Detection and segmentation of hyperdense middle cerebral artery sign on non-contrast CT using artificial intelligence

**DOI:** 10.1101/2024.07.25.24311036

**Authors:** Pyeong Eun Kim, Sue Young Ha, Myungjae Lee, Nakhoon Kim, Dongmin Kim, Leonard Sunwoo, Wi-Sun Ryu, Beom Joon Kim

## Abstract

**Background:** The hyperdense artery sign (HAS) in patients with large vessel occlusion (LVO) is associated with outcomes after ischemic stroke. Considering the labor-intensive nature of manual segmentation of HAS, we developed and validated an automated HAS segmentation algorithm on non-contrast brain CT (NCCT) images using a multicenter dataset with independent annotations by two experts.

**Methods:** For the training dataset, we included patients with ischemic stroke undergoing concurrent NCCT and CT angiography between May 2011 and December 2022 from six stroke centers. The model was externally validated using a dataset from one stroke center. For the clinical validation dataset, a consecutive series of patients admitted within 24 hours of symptom onset were included between December 2020 and April 2023 from six stroke centers. The model was trained using a 2D U-Net algorithm with manual segmentation by two experts. We constructed models trained on datasets annotated individually by each expert, and an ensemble model using shuffled annotations from both experts. The performance of the models was compared using area under the receiver operating characteristics curve (AUROC), sensitivity, and specificity.

**Results:** A total of 673, 365, and 774 patients were included in the training, external validation, and clinical validation datasets, respectively, with mean (SD) ages of 68.8 (13.2), 67.6 (13.4), and 68.8 (13.6) years and male frequencies of 55.0%, 59.5%, and 57.6%. The ensemble model achieved higher AUROC and sensitivity compared to the models trained on annotations from a single expert in the external validation dataset. In the clinical validation dataset, the ensemble model exhibited an AUROC of 0.846 (95% CI, 0.819–0.871), sensitivity of 76.8% (65.1–86.1%), and specificity of 88.5% (85.9–90.8%). The predicted volume of the clot was significantly correlated with infarct volume on follow-up diffusion-weighted imaging (r=0.42; p<0.001).

**Conclusion:** Our algorithm promptly and accurately identifies clot signs, facilitating the screening of potential patients who may require intervention.

## Introduction

The hyperdense middle cerebral artery sign (HAS) on non-contrast computed tomography (NCCT), also known as the clot sign, indicates thromboembolic occlusions in a vessel and is one of the earliest indicators of large vessel occlusion (LVO).^1^ This sign manifests as a high attenuation area on NCCT images, attributable to accumulated red blood cells and reduced blood flow.^2^ The volume and length of HAS have been associated with infarct swelling,^3^ successful recanalization rates,^4^ and clinical outcomes in patients with LVO.^5^ However, accurately and rapidly measuring clot volumes through manual segmentation in clinical practice remains challenging.

Manual clot segmentation, although the standard procedure in clinical research for image-based clot analysis,^1^ is increasingly impractical due to its labor-intensive nature and the difficulty in consistently and accurately delineating thrombus regions. To address these challenges and facilitate volumetric clot measurement in large-scale studies, several automated clot segmentation methods employing artificial intelligence (AI) have been developed.^6–8^ Despite their promising results, these methods face limitations, including a lack of robust external validation^6,7^ and restricted application to thin-section NCCT.^8^ Furthermore, moderate inter-expert agreement on the presence of HAS^7,9^ suggests that training an AI model on a diverse dataset with annotations from multiple experts could enhance its accuracy, an area yet to be fully explored.

This study leveraged NCCT and concurrent CT angiography (CTA) scans from seven comprehensive stroke centers (n=1,038) to accurately delineate the presence of HAS, which are challenging to identify on thick-section (≥3mm) NCCT alone. Using this data, we developed a robust HAS segmentation algorithm and examined the inherent variability in HAS identification through independent annotations from experienced experts. Additionally, we explored the clinical implications of our algorithm by analyzing correlations between the predicted length and volume of HAS and patient outcomes, specifically focusing on infarct volume on diffusion-weighted imaging (DWI) and functional outcomes, using a clinical dataset (n=774) from six comprehensive stroke centers.

## Materials and Methods

### Study population

For training and internal validation datasets, we retrospectively collected data of patients with ischemic stroke (admitted between May-2011 and December-2022) across six comprehensive stroke centers, using the following criteria: (1) age≥18 years; (2) admission within 7 days of symptom onset; and (3) concurrent NCCT and CTA acquisition within 10 min interval between exams.

For the external validation dataset, patients with ischemic stroke (admitted between July-2010 and November-2022) were retrospectively collected from one comprehensive stroke center, adhering to the aforementioned criteria.

To clinically validate our algorithm, we retrospectively collected a consecutive series of 941 patients with ischemic stroke or transient ischemic attack who were not overlapped with training and internal validation, and external validation datasets between December-2020 and April-2023 from six comprehensive stroke centers using the following criteria: (1) age≥18 years; (2) admission within 24 hours of symptom onset; and (3) concurrent NCCT and CTA acquisition within 10 min interval between exams.

In all cases, patients underwent both NCCT and CTA concurrently to evaluate LVO at the discretion of attending physicians. Patients were excluded based on the following criteria: (1) presence of brain tumors, intracranial hemorrhages, or surgical implants such as ventriculoperitoneal shunts or extraventricular drainage catheters, (2) NCCT scan exhibiting residual contrast agents, (3) NCCT scan with poor image quality, and (4) bilateral LVO (Figure 1). The institutional review boards of Seoul National University Bundang hospital approved the study and waived informed consent due to the anonymous and retrospective study design. The data that support the findings of this study are available from the corresponding author upon request. The study design adheres to the STARD guidelines (Standards for Reporting of Diagnostic Accuracy Studies).^10^

**Figure 1.**
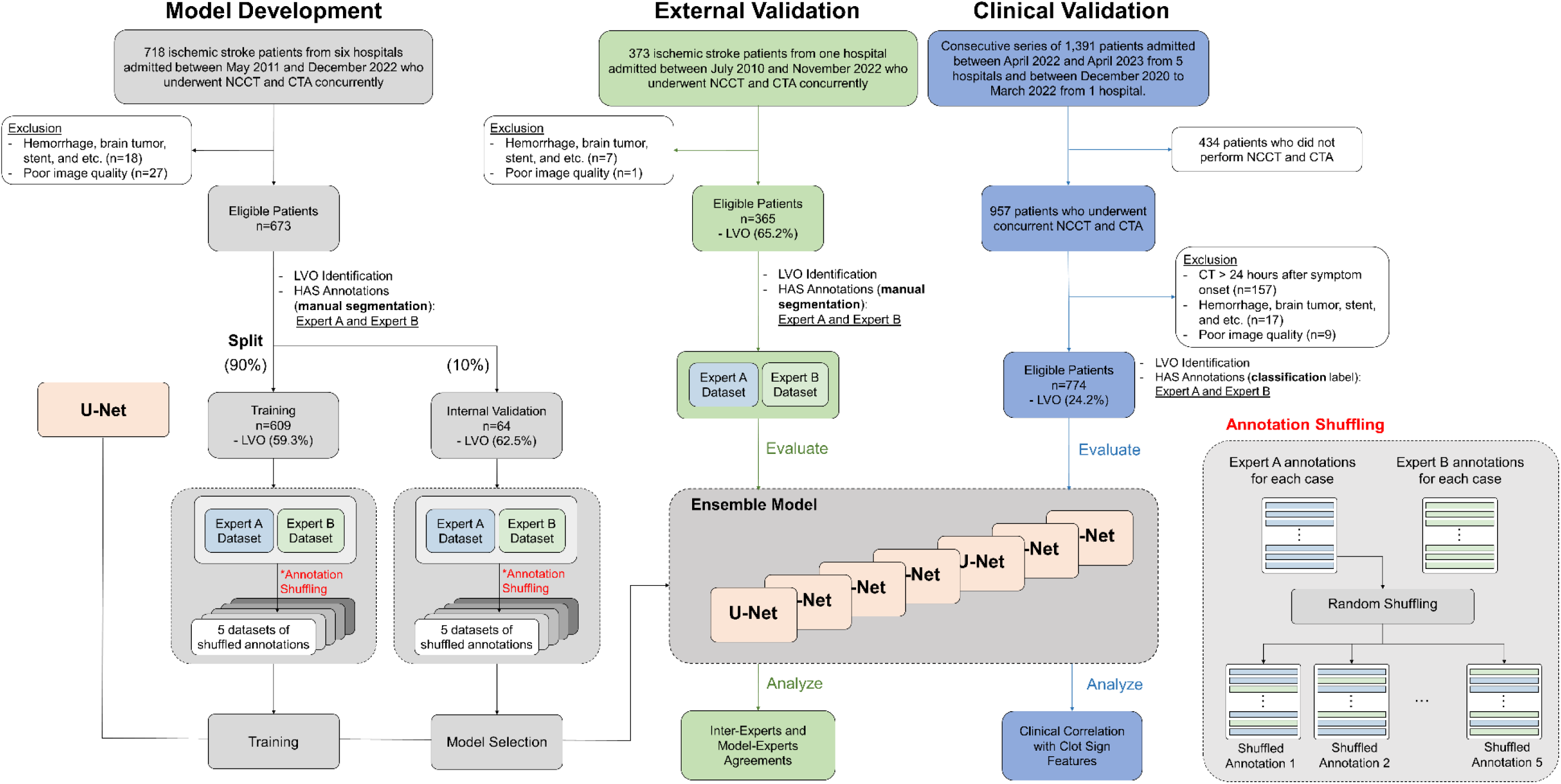
Study flow chart. NCCT=noncontrast CT; CTA=CT angiography; LVO=large vessel occlusion; HAS=hyperdense artery sign

### Clinical data collection

Using standardized protocol, demographic data, medication histories and details on vascular risk factors were collected.^11,12^ Stroke subtypes were determined by consensus among neurologists at each participating center using a validated magnetic resonance imaging (MRI)-based algorithm. Admission National Institutes of Health Stroke Scale (NIHSS) scores, pre-stroke modified Rankin Scale (mRS) scores, and 3-month mRS scores were evaluated by certified (http://www.stroke-edu.or.kr/) physicians in each hospital.

### Hyperdense artery sign annotations

Identification of HAS on thick-section NCCT scan alone is prone to misdiagnosis due to calcification, partial volume averaging, high hematocrit value, or cases with spontaneous recanalization.^13^ Thus, only patients who are confirmed to have LVO based on concurrent CTA were subjected to HAS annotation. Scans were adjusted to a window width of 80 HU and a center level of 35-40 HU. The presence of HAS was determined visually, meaning the decision was based solely on visual assessment without using any Hounsfield unit (HU) threshold as previously described.^4^ Manual segmentation of clots on NCCT was performed independently by two experts (with 8 and 20 years of experience, respectively) without referencing CTA. This approach ensured diversity in the training data by incorporating the perspectives of different experts.

### Identification of LVO

In this study, we defined LVO as a complete contrast filling defect in intracranial carotid artery or middle cerebral artery M1/M2 segment. The presence, side, and location of LVO was determined referring to CTA which was concurrently performed with NCCT by an experienced neurologist. Subsequently, these diagnoses were compared with the stroke registry data, which have been independently verified by vascular neurologists at each comprehensive stroke center. A consensus was made if discordance in diagnosis was found. All data were analyzed in a central image laboratory.

### Data preparation

In the training and validation dataset, patients from six stroke centers were randomly split in a 9:1 ratio into training and internal validation datasets. HAS segmentation masks were independently obtained by two experts. To simulate the inherent variability in HAS identification on NCCT scans, a new set of labels was created by randomly mixing the annotations from the two experts (Figure 1). To be specific, given a dataset of NCCT scans with corresponding annotations from both expert 1 and expert 2, a new set of clot sign segmentation masks was generated by randomly selecting annotations from either expert 1 or 2 for each case. To comprehensively capture the variability and potential disagreement between the experts, five additional sets of HAS annotations were constructed using this random selection process. This resulted in a total of seven sets of annotations, including the two original annotations from both experts.

### Deep Learning Segmentation Model Training and Evaluation

We trained a deep learning-based semantic segmentation model using a combination of Dice loss and binary cross-entropy loss functions. The model architecture was based on a 2D U-Net^14^ with modifications to the number of down-sampling stages, feature dimensions, and normalization methods. As input, three consecutive slices of NCCT scan were windowed (width:50, level:60), resized to 256×256 pixels, normalized, and stacked into a three-channel image. The model outputs pixel-wise probabilities of HAS.

To prevent overfitting and maximize data variability, we applied various data augmentation techniques, including rotation, horizontal flipping, translation, brightness adjustment, zooming, noise addition, and random erasing. Detailed information on the model structure and training procedures is provided in the Supplemental Material (Figure S1).

We developed three different deep learning models in this study. Model 1 and Model 2 were trained solely using the data annotated by Expert 1 and Expert 2, respectively. To develop an ensemble model trained on data annotated by different experts, we trained the model on each set of NCCT scans and their corresponding annotations, resulting in seven distinct models. Model selection was performed using the internal validation dataset based on the Dice similarity coefficient (DSC). An ensemble method was then employed by averaging all pixel-wise predictions from the seven models. The performance of the ensemble model was compared to the models trained on single datasets annotated by each expert, referred to as Model 1 and Model 2, respectively. The metrics used to evaluate the models are as follows (detailed definitions are provided in the Table S1):

- Volume-based metrics: Spearman’s correlation for volume and absolute volume difference (AVD) [mm^3^]
- Overlap metrics: DSC
- Diagnostic metrics: sensitivity, specificity, positive predictive value (PPV), negative predictive value (NPV), and predicted pixel-based area under the receiver operating characteristic curve (AUROC)

Additionally, to evaluate the robustness of the model’s performance and the inherent variability between experts, we assessed the consistency of model predictions on the internal and external validation datasets with manual segmentations by the two experts. For this analysis, only patients confirmed to have LVO were included, as no annotations were performed on patients without LVO. In cases where the prediction of HAS was correct but the location was incorrect (DSC=0), the case was designated as a false negative.

To validate the feasibility of our algorithm for use in a clinical setting, we conducted an analysis of its processing time. We randomly selected 100 NCCT scans, each with a slice thickness of 5mm, from the clinical validation dataset. The total processing time measured encompasses stages of reading DICOM files, pre-processing, and post-processing. The test was conducted on limited hardware resources (VRAM of 2GB and 8 CPU cores).

### Clinical validation dataset assessment

In the clinical validation dataset, the presence of LVO was determined by an expert. NCCT scans were independently reviewed by two experts, and the presence of HAS was defined when both experts agreed. The length of the clot predicted by the algorithm was defined as the maximal diameter of the clot in the 2D plane. Infarct volume on DWI was automatically calculated using validated software (JLK-DWI, JLK Inc., Seoul, Korea)^15,16^ and the segmented infarcts were meticulously reviewed by an expert to ensure correct segmentation. Information on the endovascular procedure including modified Thrombolysis in cerebral infarction (mTICI) scale after endovascular treatment (EVT), and 3-month modified Rankin scale (mRS) were retrieved from a prospective stroke registry.

### Statistical Analysis

To compare baseline characteristics between the training, internal, and external validation datasets, we used ANOVA or the Kruskal-Wallis test for continuous variables, and the chi-square test or Fisher’s exact test for categorical variables, as appropriate. For comparisons of inter-expert agreement with model-expert agreements, we employed the paired t-test of the mean metric values. Confidence intervals for the evaluation metrics were calculated using bootstrapping with 1000 iterations. In this analysis, the number of predicted voxels was used as the criterion. Spearman’s correlation analysis was used to assess the clot volume correlation between manual and automated segmentations. In the clinical validation dataset, the frequency of HAS between stroke subtypes was compared using the chi-square test. Associations of the predicted clot length and volume with procedure time in patients undergoing endovascular procedures were analyzed using Pearson’s correlation analysis. Additionally, patients undergoing the procedure were stratified into two groups based on post-procedure mTICI scores (0, 1, and 2a versus 2b and 3), and length and volume were compared using the t-test. The association of predicted clot length or volume with infarct volumes on DWI was analyzed using Pearson’s correlation analysis. Furthermore, clot length and volume were compared by 3-month mRS scores using ANOVA. All statistical analyses were performed using STATA 16.0 (STATA Corp., Texas, USA), and a p<0.05 was considered statistically significant.

## Results

### Baseline characteristics

Following the exclusion criteria, 45, 8, and 172 patients were excluded from training and validation, external validation, and clinical validation datasets (Figure 1), respectively, remaining 673, 365, and 774 patients. The mean±SD ages were 68.8±13.2, 67.6±13.4, and 68.8±13.6 years and frequencies of male were 55.0%, 59.5%, and 57.6%, respectively (Table 1). LVO was more frequent in external validation datasets than other datasets (59.4%, 65.2%, and 24.2% in training and validation dataset, external validation dataset, and clinical validation dataset, respectively; p<0.0001). Accordingly, HAS were more frequent in the external validation dataset regardless of which experts performed the labeling (p=0.038). CT vendors and imaging parameters were significantly different between datasets, indicating heterogeneity of datasets. Slice thickness of NCCT scans ranged from 3.0 to 5.0mm.

**Table 1.**
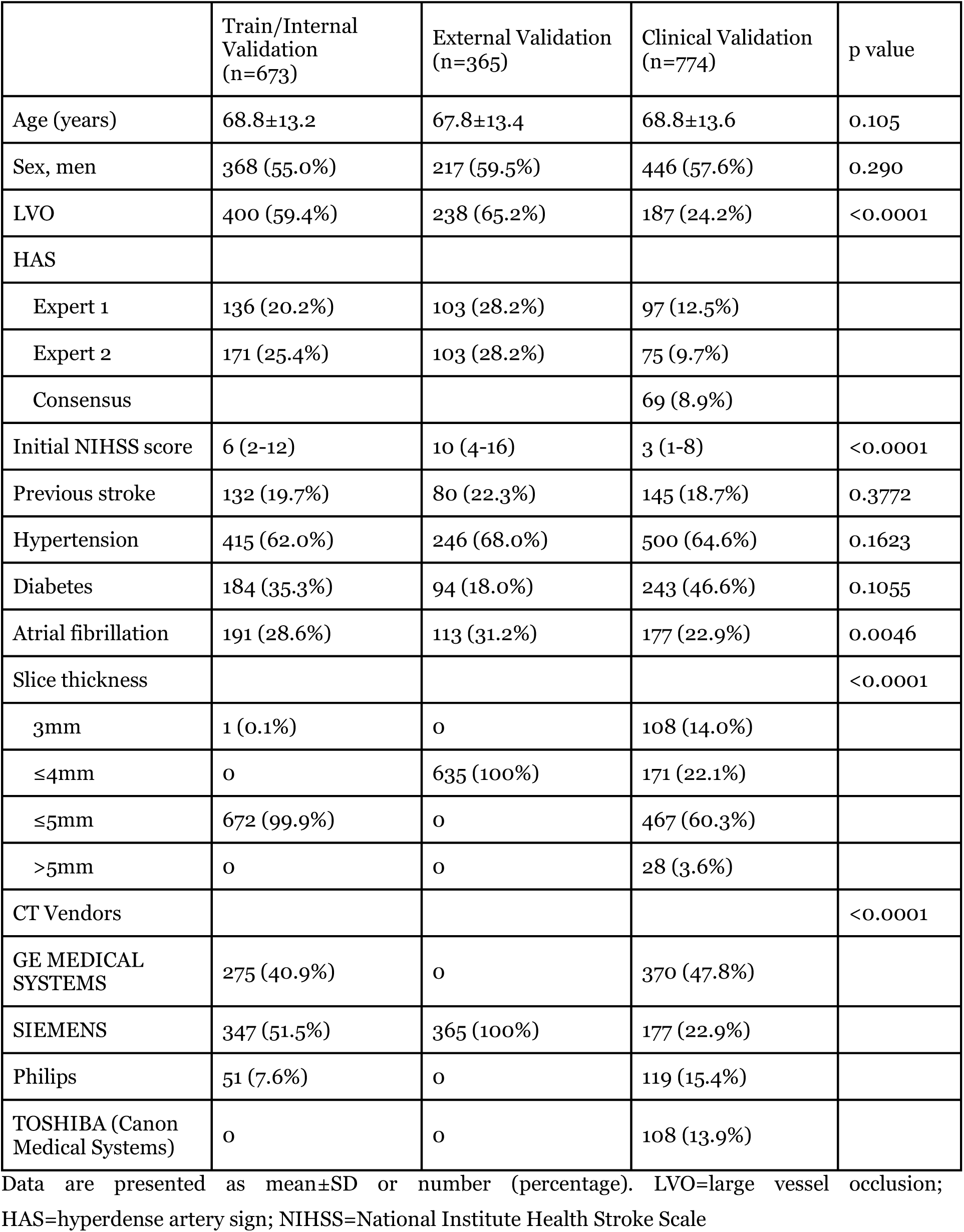
Baseline characteristics of study population.

|  | Train/Internal Validation<br>(n=673) | External Validation<br>(n=365) | Clinical Validation<br>(n=774) | p value |
| --- | --- | --- | --- | --- |
| Age (years) | 68.8±13.2 | 67.8±13.4 | 68.8±13.6 | 0.105 |
| Sex, men | 368 (55.0%) | 217 (59.5%) | 446 (57.6%) | 0.290 |
| LVO | 400 (59.4%) | 238 (65.2%) | 187 (24.2%) | <0.0001 |
| HAS |  |  |  |  |
| Expert 1 | 136 (20.2%) | 103 (28.2%) | 97 (12.5%) |  |
| Expert 2 | 171 (25.4%) | 103 (28.2%) | 75 (9.7%) |  |
| Consensus |  |  | 69 (8.9%) |  |
| Initial NIHSS score | 6 (2-12) | 10 (4-16) | 3 (1-8) | <0.0001 |
| Previous stroke | 132 (19.7%) | 80 (22.3%) | 145 (18.7%) | 0.3772 |
| Hypertension | 415 (62.0%) | 246 (68.0%) | 500 (64.6%) | 0.1623 |
| Diabetes | 184 (35.3%) | 94 (18.0%) | 243 (46.6%) | 0.1055 |
| Atrial fibrillation | 191 (28.6%) | 113 (31.2%) | 177 (22.9%) | 0.0046 |
| Slice thickness |  |  |  | <0.0001 |
| 3mm | 1 (0.1%) | 0 | 108 (14.0%) |  |
| ≤4mm | 0 | 635 (100%) | 171 (22.1%) |  |
| ≤5mm | 672 (99.9%) | 0 | 467 (60.3%) |  |
| >5mm | 0 | 0 | 28 (3.6%) |  |
| CT Vendors |  |  |  | <0.0001 |
| GE MEDICAL SYSTEMS | 275 (40.9%) | 0 | 370 (47.8%) |  |
| SIEMENS | 347 (51.5%) | 365 (100%) | 177 (22.9%) |  |
| Philips | 51 (7.6%) | 0 | 119 (15.4%) |  |
| TOSHIBA (Canon Medical Systems) | 0 | 0 | 108 (13.9%) |  |

Data are presented as mean±SD or number (percentage). LVO=large vessel occlusion; HAS=hyperdense artery sign; NIHSS=National Institute Health Stroke Scale

### Performance of deep learning models detecting and segmenting hyperdense artery sign

In the internal validation dataset, the models showed sensitivity of 0.630~0.877, specificity of 0.702~0.835, and AUROC of 0.727~0.889. In the external validation dataset, the models showed sensitivity of 0.729~0.817, specificity of 0.595~0.826, and AUROC of 0.755~0.839. For both datasets, the ensemble model showed a slightly higher AUROC compared to the other two models (Table 2).

**Table 2.**
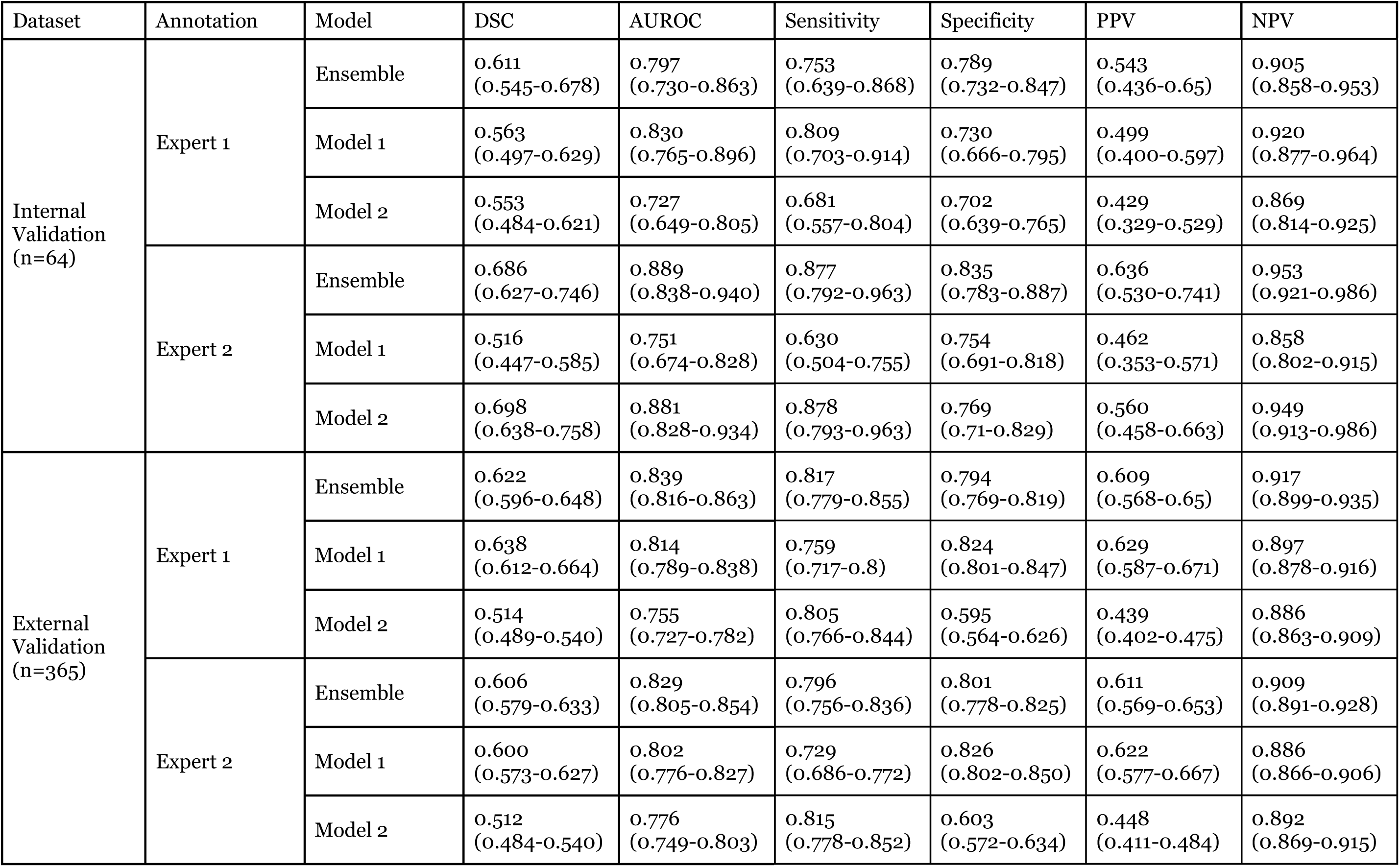
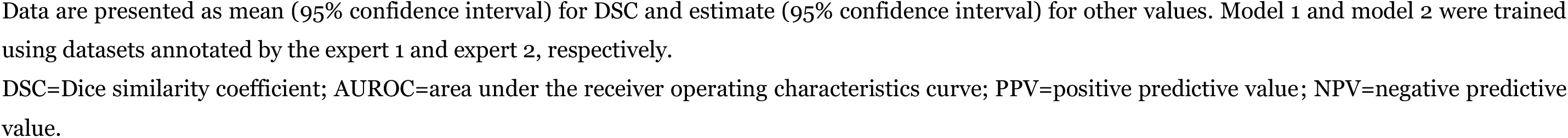
Performance of deep learning models in internal and external validation datasets.

| Dataset | Annotation | Model | DSC | AUROC | Sensitivity | Specificity | PPV | NPV |
| --- | --- | --- | --- | --- | --- | --- | --- | --- |
| Internal Validation<br>(n=64) | Expert 1 | Ensemble | 0.611<br>(0.545-0.678) | 0.797<br>(0.730-0.863) | 0.753<br>(0.639-0.868) | 0.789<br>(0.732-0.847) | 0.543<br>(0.436-0.65) | 0.905<br>(0.858-0.953) |
|  |  | Model 1 | 0.563<br>(0.497-0.629) | 0.830<br>(0.765-0.896) | 0.809<br>(0.703-0.914) | 0.730<br>(0.666-0.795) | 0.499<br>(0.400-0.597) | 0.920<br>(0.877-0.964) |
|  |  | Model 2 | 0.553<br>(0.484-0.621) | 0.727<br>(0.649-0.805) | 0.681<br>(0.557-0.804) | 0.702<br>(0.639-0.765) | 0.429<br>(0.329-0.529) | 0.869<br>(0.814-0.925) |
|  | Expert 2 | Ensemble | 0.686<br>(0.627-0.746) | 0.889<br>(0.838-0.940) | 0.877<br>(0.792-0.963) | 0.835<br>(0.783-0.887) | 0.636<br>(0.530-0.741) | 0.953<br>(0.921-0.986) |
|  |  | Model 1 | 0.516<br>(0.447-0.585) | 0.751<br>(0.674-0.828) | 0.630<br>(0.504-0.755) | 0.754<br>(0.691-0.818) | 0.462<br>(0.353-0.571) | 0.858<br>(0.802-0.915) |
|  |  | Model 2 | 0.698<br>(0.638-0.758) | 0.881<br>(0.828-0.934) | 0.878<br>(0.793-0.963) | 0.769<br>(0.71-0.829) | 0.560<br>(0.458-0.663) | 0.949<br>(0.913-0.986) |
| External Validation<br>(n=365) | Expert 1 | Ensemble | 0.622<br>(0.596-0.648) | 0.839<br>(0.816-0.863) | 0.817<br>(0.779-0.855) | 0.794<br>(0.769-0.819) | 0.609<br>(0.568-0.65) | 0.917<br>(0.899-0.935) |
|  |  | Model 1 | 0.638<br>(0.612-0.664) | 0.814<br>(0.789-0.838) | 0.759<br>(0.717-0.8) | 0.824<br>(0.801-0.847) | 0.629<br>(0.587-0.671) | 0.897<br>(0.878-0.916) |
|  |  | Model 2 | 0.514<br>(0.489-0.540) | 0.755<br>(0.727-0.782) | 0.805<br>(0.766-0.844) | 0.595<br>(0.564-0.626) | 0.439<br>(0.402-0.475) | 0.886<br>(0.863-0.909) |
|  | Expert 2 | Ensemble | 0.606<br>(0.579-0.633) | 0.829<br>(0.805-0.854) | 0.796<br>(0.756-0.836) | 0.801<br>(0.778-0.825) | 0.611<br>(0.569-0.653) | 0.909<br>(0.891-0.928) |
|  |  | Model 1 | 0.600<br>(0.573-0.627) | 0.802<br>(0.776-0.827) | 0.729<br>(0.686-0.772) | 0.826<br>(0.802-0.850) | 0.622<br>(0.577-0.667) | 0.886<br>(0.866-0.906) |
|  |  | Model 2 | 0.512<br>(0.484-0.540) | 0.776<br>(0.749-0.803) | 0.815<br>(0.778-0.852) | 0.603<br>(0.572-0.634) | 0.448<br>(0.411-0.484) | 0.892<br>(0.869-0.915) |

Data are presented as mean (95% confidence interval) for DSC and estimate (95% confidence interval) for other values. Model 1 and model 2 were trained using datasets annotated by the expert 1 and expert 2, respectively.
DSC=Dice similarity coefficient; AUROC=area under the receiver operating characteristics curve; PPV=positive predictive value; NPV=negative predictive value.

In the internal validation dataset annotated by expert 1, the ensemble model achieved a DSC of 0.611 (95% CI, 0.545−0.678), which was comparable to Model 1 and Model 2 (p>0.05). In the dataset annotated by expert 2, the ensemble model achieved a DSC of 0.686 (0.627−0.746), which was higher than Model 1 (p<0.001) and comparable to Model 2 (p>0.05). Similar results were observed in the external validation dataset, with the ensemble model achieving higher or comparable DSCs compared to Model 1 and Model 2. In the external validation dataset, DSCs between the ensemble model and each expert (0.622 [0.596−0.648] for expert 1 and 0.606 [0.579−0.633] for expert 2) were comparable to the inter-expert DSC (p > 0.05).

The volumetric correlation between the ensemble model’s outputs and expert annotations was higher than the inter-expert correlation (0.573), with model-expert correlations of 0.673 for expert 1 and 0.627 for expert 2 (Figure 2). The AVD between the ensemble model and each expert was lower than the AVD between the experts, with model-expert differences of 45.78 (±66.01) mm³ and 54.21 (±79.38) mm³ for experts 1 and 2, respectively. The mean processing time for segmenting HAS using the algorithm was 3.18 ± 0.12 seconds on NCCT scans with 4mm thickness (n=100).

**Figure 2.**
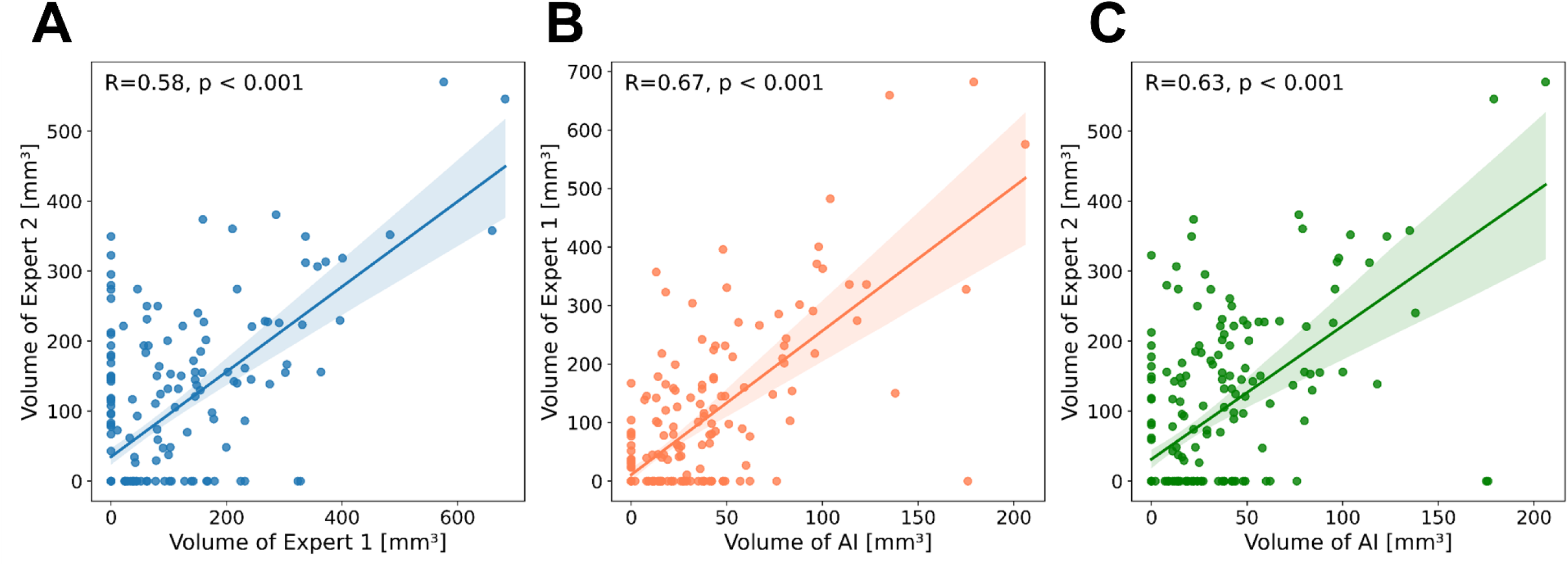
Volumetric analysis comparing clot volumes from manual annotation and automated segmentation algorithm. Dot plots for clot volumes by two experts (A), expert 1 and the algorithm (B), and expert 2 and the algorithm (C). Lines and shaded areas indicate regression lines and their 95% confidence intervals

### Inter-expert agreement on hyperdense artery sign

In the external validation dataset, Cohen’s kappa and percent agreement for the presence of HAS between experts were 0.521 and 76.5%, respectively. The DSC between experts for HAS segmentation in patients marked as HAS-positive by both experts was 0.622 (95% CI, 0.594−0.650). For patients marked as HAS-positive by either or both experts, the DSC between experts was 0.312±0.314. Spearman’s correlation coefficient and AVD for manually segmented clot volumes between experts were 0.573 and 57.09(±78.99) mm^3^, respectively.

### Review of false positive and false negative cases by deep learning algorithm

In the external validation dataset, the algorithm detected 71 false positive HAS. Among these, 24 showed symmetrically increased density in the bilateral MCAs, 24 displayed asymmetric density between the bilateral MCAs, 16 had calcified M1-MCA segments, 5 had relatively high attenuated MCAs due to adjacent chronic ischemic lesions, and 2 exhibited cortical high attenuated lesions (Figure 3). Additionally, the algorithm missed HAS in 27 patients. Among these, 11 had ambiguous density differences between the bilateral MCAs, 9 had HAS in the MCA-M2 or MCA-M3 segments, 3 had HAS in the calcified MCA, and 4 had short segment clot signs in the MCA.

**Figure 3.**
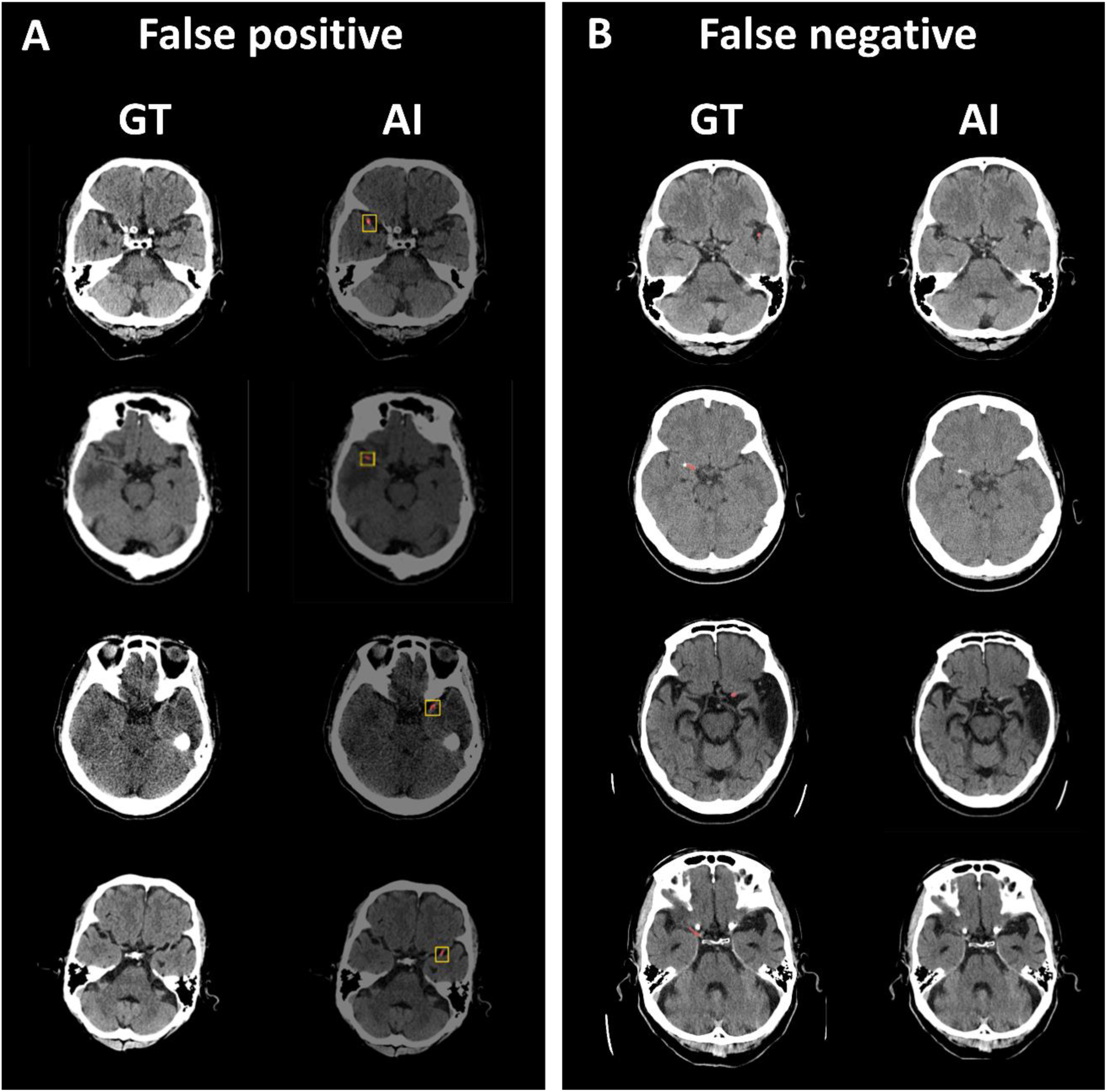
Representative false positive and false negative cases in the external validation dataset. (A) The artificial intelligence (AI) model detected calcification of the middle cerebral artery (MCA) (the first panel), high attenuation adjacent chronic lesion (the second panel), high attenuation area in the cortex (the third panel), and asymmetric density between the bilateral MCAs (the fourth panel) as hyperdense artery sign (HAS). (B) The algorithm missed HAS at the M2-MCA segment (the first panel), in the calcified MCA (the second panel), at short segment of the MCA (the third panel), and ambiguous density differences between the bilateral MCAs (the fourth panel).

### Clinical validation of clot segmentation model

In the clinical validation dataset (n=774), 187 (24.2%) patients had LVO, and 69 patients (8.9%) had a HAS. The AUROC, sensitivity, specificity, PPV, and NPV of the algorithm were 0.846 (95% CI, 0.819–0.871), 0.768 (0.651–0.861), 0.885 (0.859–0.908), 0.396 (0.339–0.455), and 0.975 (0.962–0.984), respectively. The median length and volume (interquartile range) of automatically segmented clots were 8.2 mm (5.6−17.1 mm) and 116.8 mm³ (74.4−201.1 mm³), respectively. When patients (n=118) undergoing EVT were divided into two groups (mTICI 0, 1, or 2a vs. 2b or 3), both the length and volume of the clot tended to be lower in the latter group compared with the former group, although the differences were not statistically significant (p=0.57 and p=0.19, respectively; Figure S2). In patients receiving EVT, no association was found between algorithm-detected clot length or volume and the procedure time of EVT (Figure S3).

In patients with follow-up DWI (n=702), infarct volumes on DWI were higher in the HAS-positive group than in the HAS-negative group (median [IQR] 6.98 [0.43−75.01] mL vs. 1.16 [0.21−6.87] mL, p<0.001). Additionally, the predicted length and volume of the clot were significantly correlated with infarct volume (r=0.34 and 0.42, respectively; both p<0.001; Figure 4A and 4B). When we restricted the analysis to patients with LVO (n=171), similar associations were observed (r=0.31 and r=0.36, respectively; both p<0.001; Figure 4C and 4D). Regarding the mRS score at 3 months after stroke, both the predicted length and volume of the clot tended to increase as the mRS score increased in the entire population (Figure S4). In patients with LVO, we observed a trend of increased predicted length and volume of the clot, although the difference was not statistically significant.

**Figure 4.**
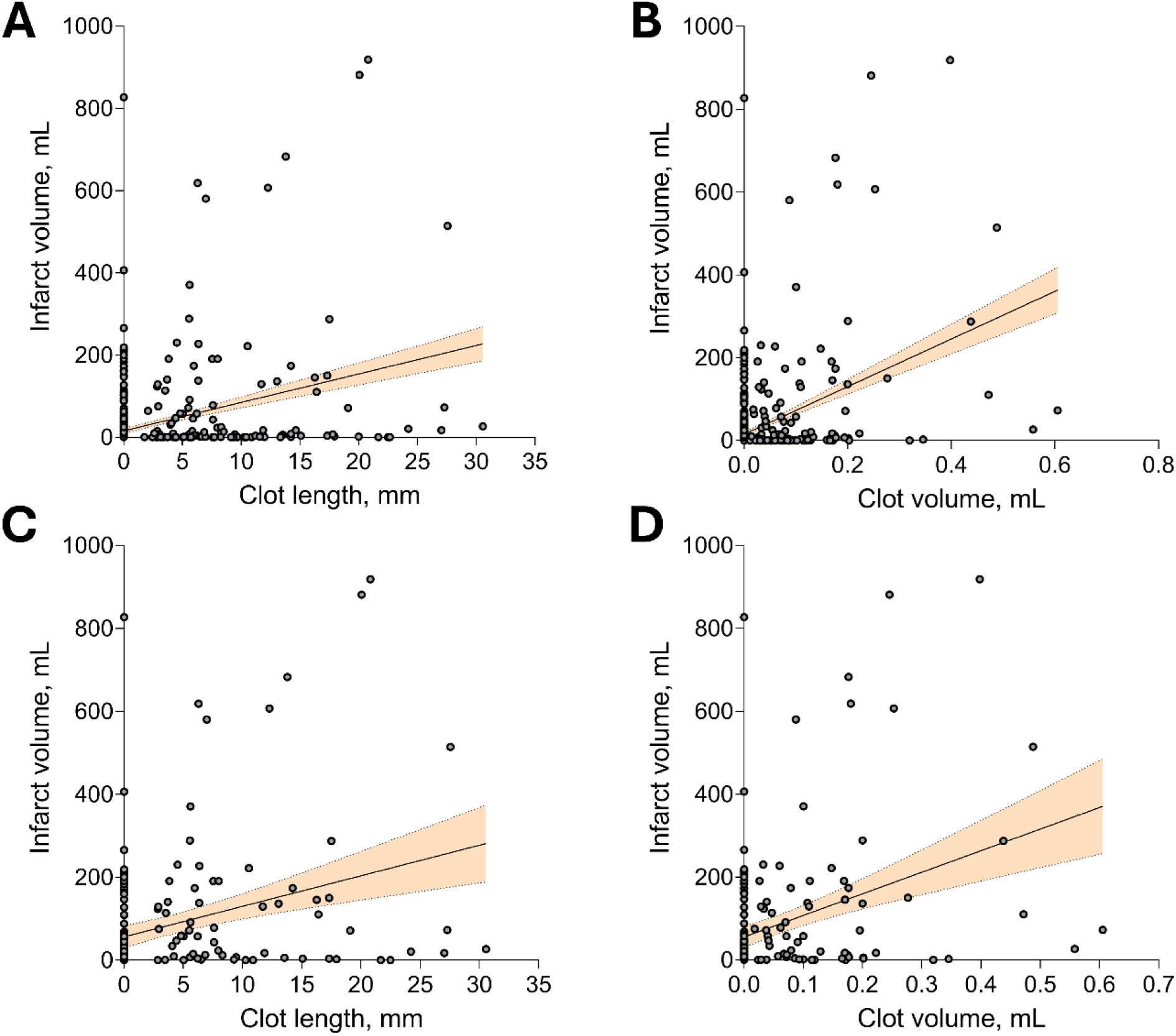
Correlation of length and volume of segmented clot and infarct volumes on follow-up diffusion-weighted image (DWI) in the clinical validation dataset. Correlation of clot length (A) and volume (B) with infarct volume on DWI in all patients. Correlation of clot length (C) and volume (D) with infarct volume on DWI in patients with large vessel occlusion. Lines and shaded areas indicate regression lines and their 95% confidence interval.

## Discussion

In the present study, we trained the deep learning model segmenting HAS using independently annotated data from two experts in patients with LVO confirmed by CTA. The model demonstrated stable performance for both datasets annotated by different experts. In the clinical validation, the model effectively detected HAS and predicted length and volume of HAS were associated with infarct volume on DWI and functional outcomes. To the best of our knowledge, our algorithm was trained on the largest dataset to date. Additionally, this is the first study to present clinical validation results of automated HAS segmentation.

Several observational studies has shown that HAS was present in 40–50% of patients with LVO.^17–19^ Previous studies utilizing AI have demonstrated promising results in detecting HAS.^4,6,20^ In a recent study, however, the AI software achieved a sensitivity of 77%, even though the standard reference was defined as LVO on CTA, raising concern about selection bias.^7^ Another study demonstrated an AI algorithm with 82.9% sensitivity and 89.7% specificity for detecting HAS,^6^ which is comparable to our results. However, they did not validate the algorithm in the external dataset not used in the training. In our study, we not only validated the segmentation results on an external dataset not used for training but also tested the detection performance on another clinical validation dataset that did not overlap. Additionally, the size of our training data is larger compared to previous studies. Considering that NCCT images can vary significantly depending on the filters used by different vendors, our algorithm, validated with a dataset comprising diverse CT vendors, may provide more reliable results compared to algorithms proposed in prior studies.

The inter-expert agreement for HAS in the present study was 0.521, aligning with the prior study by Mair et al.,^21^ in which the inter-observer kappa was 0.59 in 273 patients (25% having HAS). These results indicate that HAS detection is rather subjective. While previous studies trained their models using annotation data from a single expert or consensus or experts which possibly biased to the arbiter,^7,8,22,23^ we trained our model using shuffled annotations from two experts. This approach makes our model more consistent and robust when applied to external datasets. This assumption is further corroborated by the consistent performance of our algorithm on the external dataset annotated by two experts, and the observation that the performance of the model trained on one expert’s annotations is compromised when validated on another expert’s data.

Despite the publication of many AI segmentation models in the medical field, few are utilized in clinical practice. This is partly due to a lack of validation not only in laboratory settings but also with clinical data based on patient outcomes.^24,25^ In our study, we compared the volume and length of automatically segmented HAS with patient outcomes using a large multicenter dataset. A study of the Third International Stroke Trial (IST-3)^3^ showed that the presence of HAS at baseline was associated with a 2.2-fold increased risk of symptomatic infarct swelling, which aligns with our observation of a positive association between the length and volume of segmented clots and infarct volume on follow-up DWI. Additionally, we found a trend of association between the length and volume of clots and 3-month mRS scores, in accordance with prior studies demonstrating the association between HAS and poor functional outcomes.^26,27^ These findings extend the clinical applicability of automated detection of HAS and clot segmentation. Furthermore, each case analysis took only 3.2 seconds, making the algorithm feasible for analyzing large-scale data. This capability will facilitate further research on HAS using extensive datasets, as recent studies have highlighted the importance of clot radiomics in patients undergoing mechanical thrombectomy.^28,29^

In external validation, false positive cases were mainly associated with increased MCA density, likely due to elevated hematocrit levels or residual contrast agents.^30^ MCA calcification also contributed to false positives.^13^ False negatives often occurred when the density difference between the bilateral MCAs was ambiguous or when clots were located in the distal MCA, which the algorithm tended to miss. Recent studies have shown that incorporating brain parenchymal information along with HAS improves LVO detection performance, and utilizing NIHSS information further enhances accuracy.^11^ Therefore, combining our HAS detection algorithm with brain parenchyma analysis and NIHSS information may help mitigate the issues of false positives and false negatives.

While our study has several strengths, including training on a large dataset, utilizing independent annotations from two experts, and conducting various performance validations, there are some limitations. Firstly, although we confirmed performance with various CT vendors used in clinical practice, it is difficult to guarantee performance with vendors not included in our external or clinical validation. Secondly, our study was trained and validated on an Asian population, which limits its applicability to other ethnic groups. Although there is no evidence suggesting that the characteristics of clots or their appearance on CT differ by ethnicity, the frequency of clot signs varies among different ethnic groups.^31,32^ Therefore, our findings need to be validated in studies involving diverse ethnic populations. Thirdly, using thin slice thickness may improve the accuracy of the algorithm.^21^ However, thin section NCCT is less feasible as shown in the present study; almost hospital utilized 4–5mm slice thickness of NCCT.

NCCT is one of the most frequently performed tests in emergency rooms. Our algorithm can promptly and accurately identify HAS, helping to screen potential patients who may require intervention. This is especially beneficial in environments with a shortage of experts and has the potential to reduce the time to intervention, thereby improving outcomes for patients with ischemic stroke.

## Disclosure

M.Lee, S.Ha, P.Kim, D.Kim, and W-S.Ryu are employees of JLK Inc., Seoul, Republic of Korea.

## Supplemental Material

Table S1

Figure S1–S4

## Data Availability

The data that support the findings of this study are available from the corresponding author upon request.

## Non-standard Abbreviations and Acronyms

HAS: hyperdense artery sign
NCCT: noncontrast computed tomography
LVO: large vessel occlusion
AI: artificial intelligence
CTA: computed tomography angiography
DWI: diffusion-weighted image
NIHSS: National Institutes of Health Stroke Scale
mTICI: modified thrombolysis in cerebral infarction
EVT: endovascular treatment
mRS: modified Rankin Scale
HU: Hounsfield unit
DSC: Dice similarity coefficient
AUROC: area under the receiver operating characteristic curve
PPV: positive predictive value
NPV: negative predictive value

